# Real-time epidemiological modelling during the COVID-19 emergency in Wales

**DOI:** 10.1101/2023.08.02.23293519

**Authors:** Mike B Gravenor, Mark Dawson, Ed Bennett, Ben Thorpe, Carla White, Alma Rahat, Daniel Archambault, Noemi Picco, Gibin Powathil, Biagio Lucini

## Abstract

The sudden outbreak of the COVID-19 pandemic presented governments, policy makers and health services with an unprecedented challenge of taking real-time decisions that could keep the disease under control with non-pharmaceutical interventions, while at the same time limit as much as possible severe consequences of a very strict lockdown. Mathematical modelling has proved to be a crucial element for informing those decisions. Here we report on the rapid development and application of the Swansea Model, a mathematical model of disease spread in real time, to inform policy decisions during the COVID-19 pandemic in Wales.

## 1. Introduction

As one of the four devolved administrations of the UK, the Welsh Government (WG) has responsibility for health policy. The scientific committee for emergencies in Wales is the Technical Advisory Cell (TAC) which worked closely with SAGE, the Scientific Advisory Group for Emergencies at the UK government level. Mathematical modelling played a crucial role in the early stages of the pandemic, identifying the extent of early spread out of Wuhan, estimating transmission rates, case fatality rates and then providing the key evidence for the first UK-wide lockdown to reduce the impact on deaths (before improved therapy and vaccines were available) and to prevent the first wave overwhelming the capacity of the National Health Service (NHS) [1]. The UK lockdown suppressed virus transmission significantly, to the point when restrictions were relaxed during summer 2020, and Wales reported only a handful of cases per day. At that point, there remained great uncertainty on the future trajectory of the epidemic; to what extent it would resurge and what policies could be used to manage the next 12 months? The Swansea Model was originally designed and developed to answer these questions.

Among the Authors, MG is a mathematical epidemiologist, with experience modelling measles, malaria, avian influenza, and mad cow disease, including advising large scale control programmes [2]. MG is a member of TAC and the UK Scientific Pandemic Influenza Modelling Group (SPI-M) at which many of the key UK COVID-19 models were presented and discussed. The results from these models were fed directly into TAC and the WG. Following the largely synchronous initial lockdown, some differences in response emerged across the UK. Although general policy remained similar, timings differed and disease dynamics were often out of synch. Additionally, the demographics of Wales include a particularly elderly population, with large areas facing considerable socio-economic challenges. These issues raised the requirement for bespoke models for Wales which could quickly be tailored to the devolved policy response. In addition to the big picture of the likely course of the epidemic in Wales, and how it might be mitigated, each health board was experiencing their own planning challenges (often at different times) and in need of modelling support for day-to-day expected burden of hospital admissions and bed occupancy.

In this environment, MG was approached by colleagues at WG and Public Health Wales (PHW) to see if a policy model could be rapidly developed and deployed. It was serendipitous that BL had already contacted MG to see if expertise from the Mathematics Department (and part of the Supercomputing Wales project) could help; specifically, a small team of research software engineers (RSEs). The involvement of the RSEs was essential to the success of this project. The time pressure that all scientific advisors were working under was tremendous. Where an academic project of this type might be expected to take place over 1 or 2 years, the urgency of the pandemic could change in a matter of days. The RSEs allowed the team to evaluate and trial existing tools where available (to save time on replicating models) and to rapidly adapt the models for bespoke conditions or scenarios, albeit by working 7 days and deep into the nights. This was the start of the ‘Swansea Team’ which provided modelling support throughout the pandemic, and has expanded to continue to provide mathematical analysis for COVID-19, influenza, Respiratory Syncytial Virus (RSV) and monkey pox. It has been our philosophy throughout to maintain as close a link as we could between three components, the fundamental epidemiology of infectious disease dynamics, the underpinning mathematical framework that allows quantitative questions to be posed on the epidemiology, and a strong link with end users in the NHS, Public Health and Government so that relevant research questions were always kept to the fore.

## 2 The model

In mathematical terms, the “Swansea Model” was based on well-established epidemiological theory, using an ‘SEIR’ framework, and code published by Davies *et al*. [3]. Populations are represented in compartments, with the flow between compartments modelled by ordinary differential equations. In a simple example, a population of *N* individuals has Susceptible (*S*), Exposed (*E*), Infectious (*I*) and Recovered (*R*) compartments, with the infection processes being ‘mass-action’ and proportional to the product *S ×I /N* times the person-to-person contact rate. If *i* distinct age classes are used, then the basic model is:

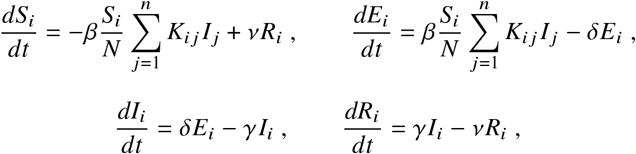

where ^−1^ is the incubation period, ^−1^ is the infectious period, ^−1^ is the duration of immunity, *β* is the transmission coefficient and *K*_*ij*_ is a matrix of mean contacts per day between each age-group. Our model employed 16 age groups, and was also expanded upon to include: time-dependent contact rates, gamma distributions for the incubation and infectious periods, and additional compartments for those individuals that were: asymptomatic infectious (*I*_*a*_, which are less infectious), pre-symptomatic infectious (*I* _*p*_), and symptomatic infectious (*I*_*s*_). Vaccination was modelled by additional compartments *S*_*u*_ and *R*_*u*_, which removed Susceptible and Recovered individuals, respectively. This transmission model was linked to a separate simple model in ’post-processing’ to keep track of clinical events, defined as delayed functions of the incidence of symptomatic infections. Thus, realistic statistical distributions could be included as delay terms to generate the incidence of clinical cases (*C*), ward hospitalisations (*H*_*w*_), intensive care unit (ICU) admissions (*H*_*ICU*_), and deaths (*D*). By including a duration distribution for each clinical event, the prevalence of cases and occupancy statistics for hospital and intensive care beds were also calculated. Clinical outcomes were essential, since almost all policy target outcomes were events such as cases (for test and trace), deaths, and hospital capacity. Partial vaccine effectiveness was included in the transmission model, as well as different vaccine effectiveness for preventing each of the clinical events. A schematic of the model is shown in Fig. 1. Lastly, we used a separate model for each of the 22 Welsh Local Authority areas, with flexible migration terms to allow for travel, and spread of infected individuals between areas. A stochastic formulation was developed, but rarely used in practice. The model was also compared to a published agent-based model (“Open ABM-COVID 19”), which generated similar results.

**Fig. 1.**
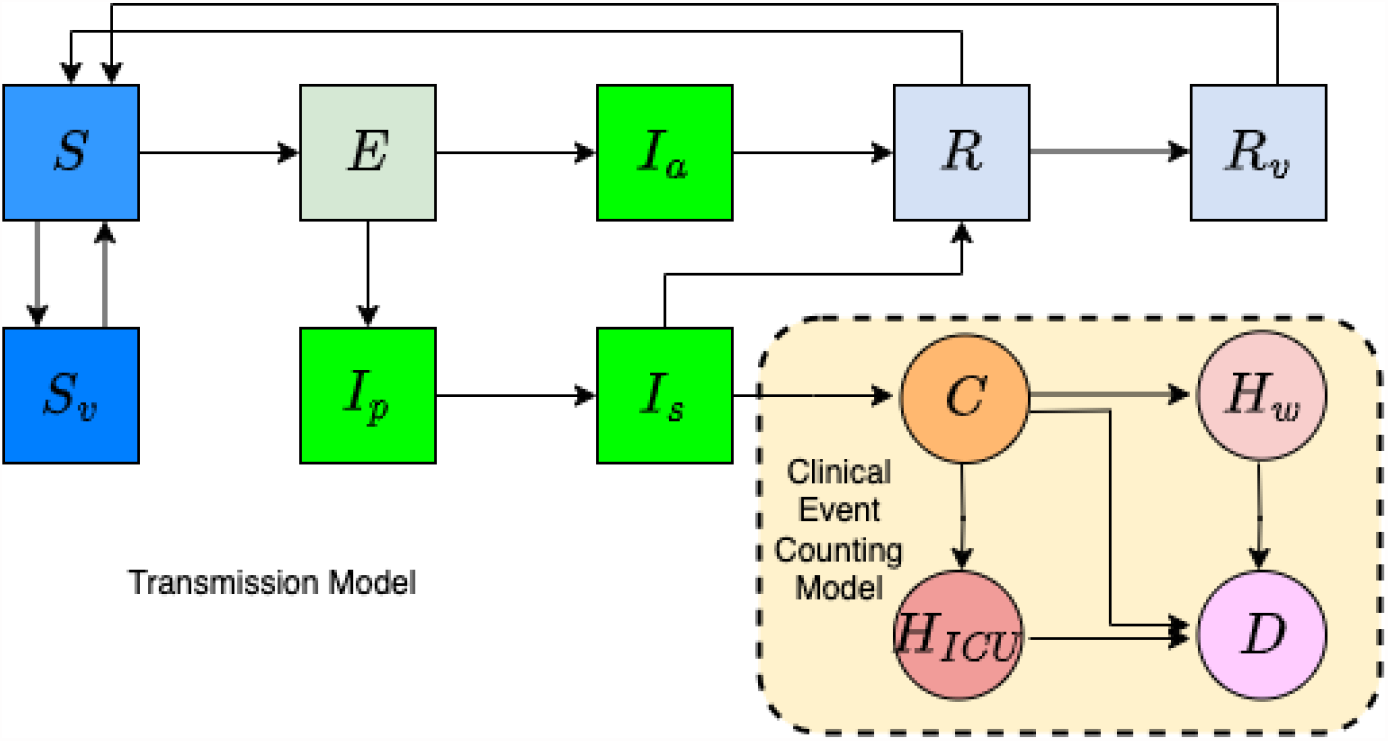
Schematic of the transmission process, and clinical event counting model.

Central to the application of the model was a ‘schedule’ of time- and age-dependent contact rates. Starting with typical contacts obtained from epidemiological surveys, the schedule employed a scaling of these contact rates which represented changes to the typical contacts of the population due to behavioural responses and imposition of non-pharmaceutical interventions (NPIs). Note that the contact rate schedule determines the widely reported effective reproduction number *R*_*t*_. When *R*_*t*_ was greater than 1 we were facing an exponentially growing epidemic, and for *R*_*t*_ less than 1 the infected incidence was declining.

The contact schedule could be applied retrospectively, by fitting the model to the previous trajectory of the epidemic in Wales, for example from a fit of the model to Public Health Wales time series data on cases, hospital events and deaths. Using Bayesian multi-objective optimisation methods we were able to provide estimates for a large number of parameters including the scaled reduction to ‘normal’ contact rates, and hence *R*_*t*_. The next step would be to apply the contact rate schedule prospectively, to simulate the potential future policy scenarios. For example, following a period of strong NPIs such as lockdown, a future scenario could include relaxing of restrictions and an increase in the contact rates. The model could then be used to simulate the future trajectory of key events under those assumptions. Similarly, during periods of alarming epidemic growth, the impact of proposed interventions could be explored by reducing contact rates by an appropriate amount in the schedule. Due to the age-structure, policy interventions could be explored in some detail by teasing apart the effects of movement, work-place, leisure, or school closures; or measures focused on reduction in contacts by the most vulnerable elderly population.

Our mathematical models were used for real time policy advice in several key areas: the estimate of *R* values; the efficacy of test and trace; assessing the need and impact of winter 2020 NPIs; advising on the likely prevalence of COVID-19 during proposed dates for Senedd elections; informing the road map out of lockdown under the impact of the vaccination roll-out and the emergence of the delta variant; modelling the expected course and impact of the delta wave; estimating the impact of laboratory testing errors on transmission; relaxing test and trace isolation requirements; and modelling emerging variants. Here, we focus on three case studies.

## 3 The Impact

### The Reasonable Worst Case

Summer 2020 was a period of great uncertainty. The first wave had been suppressed and case numbers were extremely low. With few NPIs in place *R*_*t*_ had returned to above 1, but the extent of a resurgence was unclear. We modelled the total population that remained susceptible and factors such as the impact of test and trace, in order to generate a range of winter scenarios. The resulting ‘Reasonable Worst Case’ (RWC), and ‘Most Likely Scenarios’ (MLS, based on tracking recent trends and making short-term projections) showed the considerable epidemic potential that remained, how quickly pressures could build and the winter challenges that the NHS would face. An early RWC is shown in Fig. 2. This was published at a time when the risk of any ‘second’ wave was being questioned and many doubted it could be as large as the first [4]. The sharing of the RWC across services in Wales impacted tactical (3–6 months) and operational (1–42 days) decisions across Welsh hospitals and the ambulance service, providing the basis for staff deployment for: test-trace-protect, in-hospital testing and scheduling, laboratory staffing requirements, and local and government budget allocations. The model scenarios formed part of the central assumptions for hospital bed and critical care capacity planning, elective operating schedules, and procurement decisions in health boards and the NHS Wales Informatics Service, as part of standardised planning assumptions by the third quarter of 2020. Prior warning is required for staffing critical care, and early planning for this reduces overall costs. Based on the model, the need for a 150% increase in capacity was identified and ultimately met when the large second wave was realised.

**Fig. 2.**
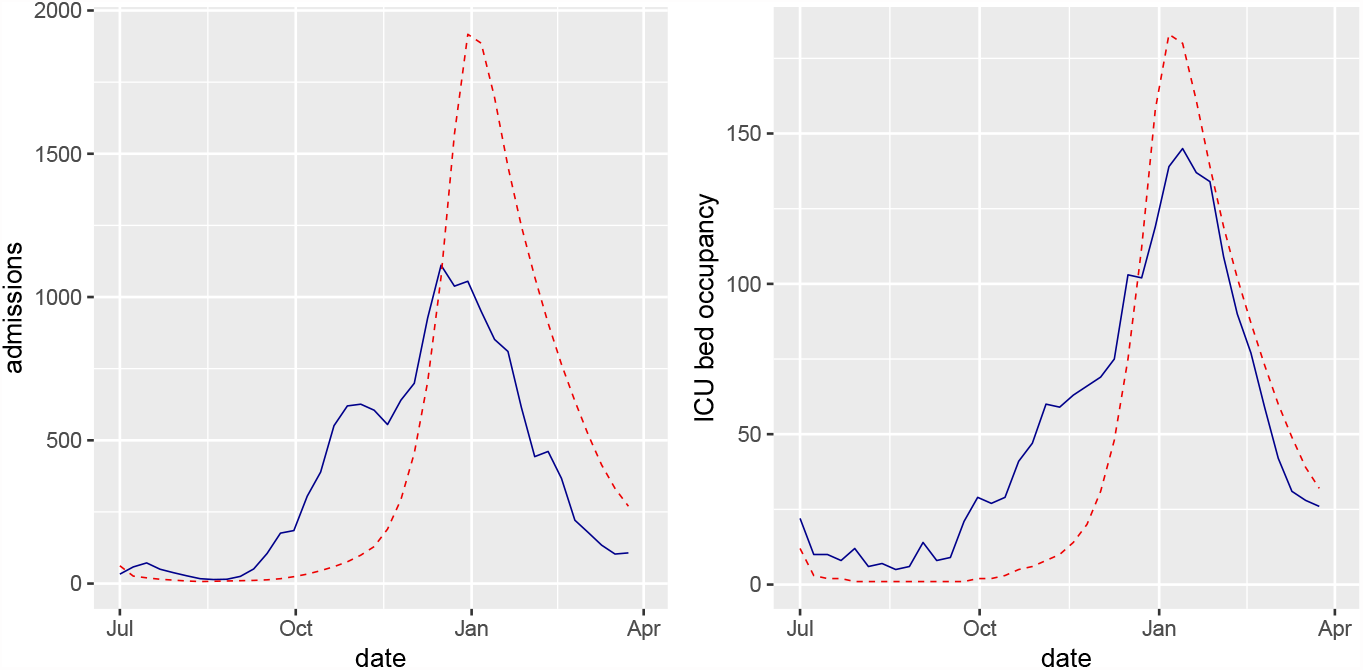
Winter 20/21 Reasonable Worst Case for weekly hospital admissions (left panel) and intensive care unit occupancy (right panel) in Wales generated in August 2020. Model scenarios are plotted in red (dashed), and the subsequent realised data in blue (solid).

### The Firebreak

By October 2020, the prevalence of COVID-19 was tracking to alarming levels. A widely discussed proposal for delaying and reducing the peak of cases was a short ‘firebreak’, through NPIs that would reduce the contacts and transmission rates. We modelled multiple different scenarios of possible policy choices coinciding with school half-terms, with different timing, level and duration of the firebreak. Each scenario was evaluated in terms of metrics such as impact on the health service, fatalities, pressure on hospital resources and the likely time ‘bought’, before case numbers would return to the previous levels after NPIs were lifted. The evidence was published simultaneously with the announcement of the 17-day Wales Firebreak on 19^th^ October 2020 [5] The impact was an effective, if short-term, intervention with an immediate reduction in *R*_*t*_ from approximately 1.4 to 0.8, resulting in a prevalence 50% lower than England, which Wales had been tracking during the autumn. There was a 6-week period before return to pre-firebreak prevalence, closely following expectations. Vital time was gained to prepare for winter seasonal peaks in hospitals, which would have been harder to manage if cases had continued their trajectory from a higher November baseline. Crucially, cases, deaths, and hospitalisations were significantly reduced (see Fig. 3).

**Fig. 3.**
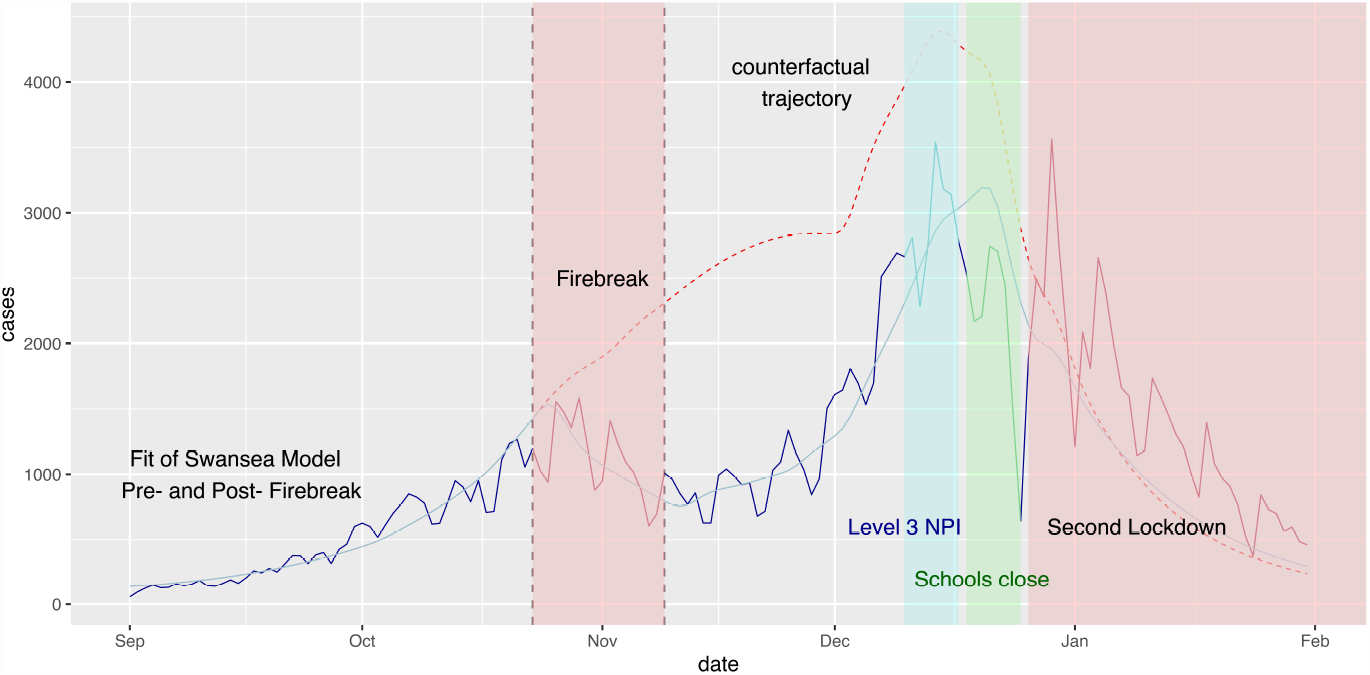
Counterfactual analysis for Firebreak, contrasting the latter to a scenario without the intervention. Subsequent NPIs were re-introduced to control the December levels of COVID-19.

### Road-map out of restrictions

Following the second major set of UK NPIs to control the second wave, the UK vaccination programme was initiated. Our efforts then focused on the exceptionally difficult interaction between the impact of the vaccination programme, and the relaxing of NPIs. At this point the models had to be expanded to include new variants, vaccination and waning immunity. A further complication was the differential effects of the vaccine, being highly effective on the more severe clinical events, but less effective at transmission blocking. In early 2021, the models showed that even after the initial dose vaccine roll out to the elderly and vulnerable population, should all NPIs have been removed at that point there remained a very large susceptible population and a high risk of burden of disease and health service impact. These models ultimately informed the staggered road-map out of NPIs, and were used early in 2021 to plan school re-opening as a priority. In January 2021, we modelled staged school re-openings, showing that there would be a small impact on serious event rates as long as other NPIs were prioritised at that time. The youngest pupils returned to face-to-face teaching on February 22^nd^ 2021 and the rest on March 15^th^.

Deeper into the vaccine roll out, the model was used to analyse the staged removal of remaining NPIs, and show the limited impact on the most severe clinical events, albeit with a very high expected prevalence of infection due to the size of the remaining susceptible population and the high transmissibility of the circulating virus variants. The combination of ’what-if’ scenarios, and short term forecasts based on the model continued to inform the analysis of the delta and omicron variant waves through to 2023, and influence the way winter planning for other seasonal viral infections such as influenza and RSV is conducted.

## Data Availability

All data produced in the present study are available upon reasonable request to the authors

## Acknowledgements

The initial year of work was conducted voluntarily and therefore underpinned by our University and Supercomputing Wales posts. Such extensive *pro bono* work highlights an important consideration for future emergency scientific advice planning. We have since had funding to continue and expand the work, and we acknowledge the European Regional Development Fund, EPSRC (grant EP/W01226X/1) and Welsh Government Grants: Sêr Cymru III – Visual Analytics of Epidemic Simulations with Many Actor Behaviours, C098/201/2022, and [SAP]4400015698. We thank Konstans Wells, Miguel Lurgi, Rhodri Fabbro, Finley Gibson and Thomas Torseny-Wier for their contributions. We are indebted to so many colleagues for support on the model and applications: members of TAC, SAGE and SPI-M; especially Brendan Collins, Craiger Solomons, Jenny Morgan, John Watkins, Andrew Nelson, James Cooke, Nick Davies, Rob Orford, Fliss Bennee, Chris Williams, Alun Lloyd, Simon Frost, Rowland Kao, Graham Medley and Laura Andrews.

## Open Access Statement

For the purpose of open access, the authors have applied a Creative Commons Attribution (CC BY) licence to any Author Accepted Manuscript version arising.

## Research Data and Code Access Statemen

Our implementation of the model is available at https://github.com/SwanMod/covid-model. The data generated for this manuscript can be requested from the authors.

